# Association between comorbidities and the risk of death in patients with COVID-19: sex-specific differences

**DOI:** 10.1101/2020.05.22.20109579

**Authors:** Mingyang Wu, Shuqiong Huang, Jun Liu, Yanling Shu, Yinbo Luo, Lulin Wang, Mingyan Li, Youjie Wang

**Affiliations:** Department of Maternal and Child Health, School of Public Health, Tongji Medical College, Huazhong University of Science and Technology, Wuhan 430030, Hubei, China.; Key Laboratory of Environment and Health, Ministry of Education & Ministry of Environmental Protection, and State Key Laboratory of Environmental Health (Incubation), School of Public Health, Tongji Medical College, Huazhong University of Science and Technology Wuhan 430030, Hubei, China.; Institute of Preventive Medicine Information, Hubei Provincial Center for Disease Control and Prevention, Wuhan 430079, Hubei, China; Central office, Qianjiang City Center for Disease Control and Prevention, Qianjiang 433199, Hubei, China; Department of Laboratory Medicine, Wuhan Children’s Hospital, Tongji Medical College, Huazhong University of Science & Technology, Wuhan 430016, Hubei, China.; Institute for Infectious Disease Control and Prevention, Hubei Provincial Center for Disease Control and Prevention, Wuhan 430079, Hubei, China.

**Keywords:** COVID-19, Diabetes, sex-specific, comorbidities

## Abstract

**Background:** The coronavirus disease 2019 (Covid-19) spreads rapidly around the world.

**Objective:** To evaluate the association between comorbidities and the risk of death in patients with COVID-19, and to further explore potential sex-specific differences.

**Methods:** We analyzed the data from 18,465 laboratory-confirmed cases that completed an epidemiological investigation in Hubei Province as of February 27, 2020. Information on death was obtained from the Infectious Disease Information System. The Cox proportional hazards model was used to estimate the association between comorbidities and the risk of death in patients with COVID-19.

**Results:** The median age for COVID-19 patients was 50.5 years. 8828(47.81%) patients were females. Severe cases accounted for 20.11% of the study population. As of March 7, 2020, a total of 919 cases deceased from COVID-19 for a fatality rate of 4.98%. Hypertension (13.87%), diabetes (5.53%), and cardiovascular and cerebrovascular diseases (CBVDs) (4.45%) were the most prevalent comorbidities, and 27.37% of patients with COVID-19 reported having at least one comorbidity. After adjustment for age, gender, address, and clinical severity, patients with hypertension (HR 1.55, 95%CI 1.35-1.78), diabetes (HR 1.35, 95%CI 1.13-1.62), CBVDs (HR 1.70, 95%CI 1.43-2.02), chronic kidney diseases (HR 2.09, 95%CI 1.47-2.98), and at least two comorbidities (HR 1.84, 95%CI 1.55-2.18) had significant increased risks of death. And the association between diabetes and the risk of death from COVID-19 was prominent in women (HR 1.69, 95%CI 1.27-2.25) than in men (HR 1.16, 95%CI 0.91-1.46) (*P* for interaction = 0.036).

**Conclusion:** Among laboratory-confirmed cases of COVID-19 in Hubei province, China, patients with hypertension, diabetes, CBVDs, chronic kidney diseases were significantly associated with increased risk of death. The association between diabetes and the risk of death tended to be stronger in women than in men. Clinicians should increase their awareness of the increased risk of death in COVID-19 patients with comorbidities.

## 1 Introduction

The number of cases of the coronavirus disease 2019 (COVID-19), caused by severe acute respiratory syndrome coronavirus 2 (SARS-CoV-2), is rising at an unprecedented rate, threatening people in various countries around the world. As of May 14, 2020, SARS-CoV-2 has affected over 4,258,666 people in at least 216 countries worldwide. ^1^ Thousands of people die of COVID-19 every day, and the number of deaths has already surpassed 300,000 globally and is expected to increase further as the disease spreads rapidly, posing a serious threat to human health.

Based on the data from WHO, the case fatality rate of SARS-CoV-2 infection varies in different regions or countries (0~19.11%). Generally, those COVID-19 cases with higher age, male sex, and more comorbidities were prone to have severe disease and subsequent mortality. ^2,3^ Most of the available studies have shown that hypertension, diabetes, and cardiovascular and cerebrovascular diseases (CBVDs) were the most prevalent comorbidities in COVID-19 patients. ^2,4,5^ In a recent study, the presence of hypertension and diabetes was respectively associated with 1.58-fold and 1.59-fold increased risk of death in patients with SARS-Cov-2 infection.^4^ Not only that, a meta-analysis of retrospective studies confirmed that COVID-19 patients with hypertension, diabetes, or CBVDs were associated with a significant increased risk of aggravation.^6^ However, because of the relatively small sample sizes in previous studies, larger sample sizes studies are needed. Moreover, sex-specific mortality of SARS-Cov-2 has been well documented, but whether there are sex-specific differences on the association of comorbidities with the risk of death in COVID-19 patients is still unknown. ^7^

Therefore, this study was conducted to 1) address the association between comorbidities and the risk of death; 2) explore the sex-specific differences on the associations of comorbidities with the risk of death in patients with COVID-19 in Hubei province (the epicenter of China).

## 2 Methods

### 2.1 Data sources and data extraction

The participants in this study were from all cases of epidemiological investigation in Hubei Province as of February 27,2020 (N=72,802). The final date of follow-up was March 7, 2020. The criteria of exclusion were as follows, 1) patients with missing value of comorbidities (n=42,011); 2) patients with age lower than 20 years (n=1288); 3) patients were diagnosed as asymptomatic or suspected or clinical diagnosed cases (n=11,038). Finally, a total of 18,465 laboratory-confirmed cases were included in the present study for analysis.

### 2.2 variables

The general characteristics and clinical features were collected using a case questionnaire at the time of diagnosis, and then imputed into the Infectious Disease Information System by local epidemiologists and public healthcare workers. The case questionnaire contains basic demographic or epidemic information (e.g., sex, birthdate, present address, occupation, and exposure history) and clinical information (e.g., symptom onset, the date of symptom onset, blood cell count, comorbidities). We computed the age of each case using the date of symptom onset and birthdate. According to the present address, all the records were further classed as residents in Wuhan and not. The clinical severity was categorized as mild, common, and severe or critical. The diagnosis criteria of clinical severity can refer to the Chinese Clinical Guidance for Covid-19 Pneumonia Diagnosis and Treatment.^8^ All the underlying comorbidities were self-reported by patients, and were classified as hypertension, diabetes, cardiovascular and cerebrovascular diseases (CBVDs), lung diseases, chronic kidney diseases, chronic liver diseases, and other comorbidities. Information on death was obtained from the Infectious Disease Information System. All the confirmed cases were diagnosed based on clinical signs or symptoms as well as positive viral nucleic acid test results on throat swab samples.^9^ Waiver of informed consent for collection of epidemiological data from patients with COVID-19 was granted by the National Health Commission of China as part of the infectious disease outbreak investigation. All identifiable personal information was removed for privacy protection.

### 2.3 Statistical analysis

The continuous variables were presented as medians (interquartile ranges), and the categorical variables were expressed as counts (percentages). The associations between comorbidities and the risk of death were estimated using Cox proportional hazards regression model (“survival” and “survminer” packages in R), with the hazards ratio (HR) and 95% confidence interval (95%CI) being reported. And covariates included in the Cox model were based on prior publication, including age (20-29, 30-39, 40-49, 50-59, or ≥60 years), gender (women or men), address (Wuhan or not), and clinical severity (severe or not). Since men have a greater risk of death than women, we therefore performed a subgroup analysis based on gender. A term of *comorbidities*gender* was added into the regression model to explore the potential interaction effect of gender on the association between comorbidities and the risk of death.

All data were analyzed with R software, Version 3.5.3. A two-tailed *P* value < 0.05 was considered statistically significant, and a *P* value for interaction < 0.10 was considered significant.

## 3 Results

### 3.1 Demographic and clinical characteristics

A total of 18,465 laboratory-confirmed COVID-19 cases with age higher than 20 years were included in the present study for analysis, and the epidemiological and clinical characters of the study population are shown in **Table 1**. Of these 18,465 cases, the median age was 50.53 years. 8828(47.81%) patients were females. Severe cases accounted for 20.11% of the study population. The most common onset symptoms were fever (79.52%), cough (56.42%), and fatigue (28.35%). Abnormal chest CT manifestation was identified in 82.47% of patients. As of March 7, 2020, a total of 919 cases deceased from COVID-19 for a fatality rate of 4.98% (**Table 1**).

**Table 1.**
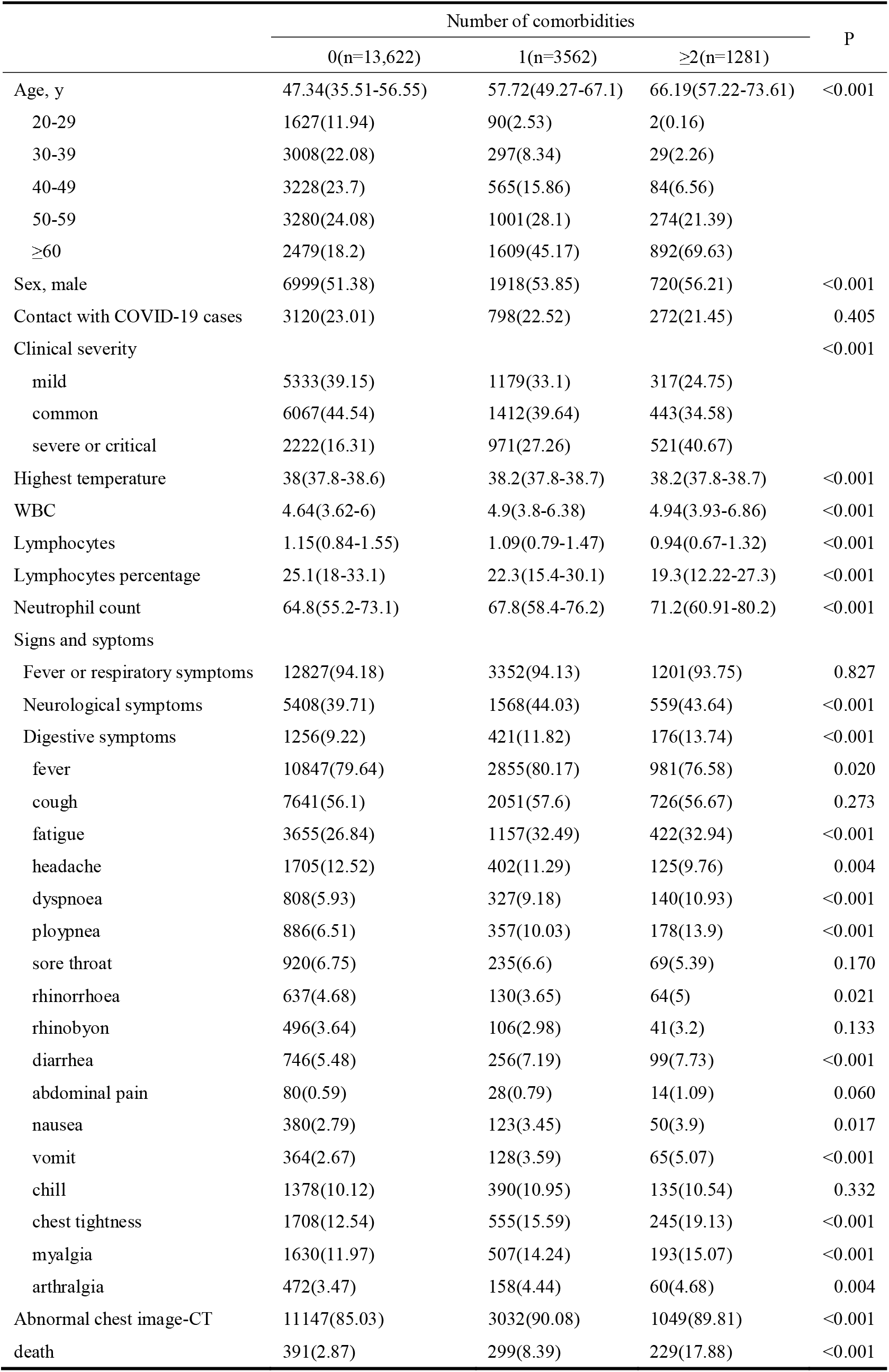
The characteristics of study population (N=18,465)

There were 3562(19.29%) and 1281(6.94%) for patients with 1 comorbidity and 2 or more comorbidities, respectively. As compared with patients without comorbidities, patients with 1 comorbidity or at least 2 comorbidities had significant higher rate of digestive symptoms (11.82% or 13.74% vs 9.22%), higher proportion of severe cases (27.26% or 40.67% vs 16.31%), and higher mortality rate (8.39% or 17.88% vs 2.87%) (**Table 1**). And the similar results were also observed in both men and women (**Table S1**).

### 3.2 The prevalence of comorbidities stratified by gender

The results of the prevalence of comorbidities in different gender are shown in **table 2**. A total of 2561(13.87%), 1022(5.53%), 822(4.45%), 463(2.51%), 149(0.81%), and 124(0.67%) patients reported having hypertension, diabetes, CBVDs, lung diseases, chronic kidney diseases, and chronic liver diseases, respectively. As compared with women, men were more likely to have at least one comorbidity (27.37% vs 24.98%). Significant higher prevalence of hypertension (14.73% vs 12.92%), lung diseases (2.92% vs 2.06%), chronic kidney diseases (0.94% vs 0.66%), and chronic liver diseases (0.84% vs 0.49%) were observed in men than in women. The prevalence of diabetes (5.81% vs 5.23%) and CBVDs (4.66% vs 4.23%) were similar in men and women (**Table 2**). Furthermore, we have further identified the epidemiological and clinical characteristics of patients with different comorbidities in men and women (**Table S2 and S3**). Of the men, patients with hypertension or diabetes or CBVDs were more likely to be older, have decreased lymphocyte or increased neutrophil counts, and have higher mortality rate. The rate of digestive symptoms in patients with hypertension was similar to that in patients without hypertension, but patients with diabetes or CBVDs had significant higher rate of digestive symptoms compared with those without diabetes or CBVDs (**Table S3**). Of the women, patients with comorbidities were more likely to be older, have decreased lymphocyte or increased neutrophil counts, have higher rate of digestive symptoms, and higher mortality rate (**Table S2**).

**Table 2.** The prevalence of comorbidities in patients with COVID-19, stratified by gender.

|  | Women | Man | P |
| --- | --- | --- | --- |
| Comorbidities | 2205(24.98) | 2638(27.37) | <0.001 |
| 0 | 6623(75.02) | 6999(72.63) | <0.001 |
| 1 | 1644(18.62) | 1918(19.9) | <0.001 |
| $\geq 2$ | 561(6.35) | 720(7.47) | <0.001 |
| hypertension | 1141(12.92) | 1420(14.73) | <0.001 |
| diabetes | 462(5.23) | 560(5.81) | 0.092 |
| CBVDs | 373(4.23) | 449(4.66) | 0.164 |
| lung diseases | 182(2.06) | 281(2.92) | <0.001 |
| chronic kidney diseases | 58(0.66) | 91(0.94) | 0.036 |
| chronic liver diseases | 43(0.49) | 81(0.84) | 0.004 |
| other comorbidities | 638(7.23) | 680(7.06) | 0.673 |

We also identified the proportion of comorbidities in women and men stratified by death status (**Table S4**). A total of 327 and 592 cases deceased from COVID-19 in women and men, respectively. Of these deceased patients, the most prevalent comorbidities were hypertension, CBVDs, and diabetes in both women and men, and the corresponding proportions of the three comorbidities were 35.47%, 18.35%, and 18.04% in women; 36.99%, 17.4%, and 13.85% in men.

### 3.3 The associations between comorbidities and death

Patients with comorbidities had significantly escalated risks of death compared with those without (**Figure S1**). In the crude model, patients with hypertension (HR 3.74, 95%CI 3.27-4.28), diabetes (HR 3.25, 95%CI 2.71-3.89), CBVDs (HR 5.08, 95%CI 4.29-6.02), lung diseases (HR 2.5, 95%CI 1.89-3.3), chronic kidney diseases (HR 5.05, 95%CI 3.55-7.18), and at least two comorbidities (HR 6.78, 95%CI 5.76-7.98) had significantly escalated risks of death than those without. After adjusting for age, the association between comorbidities and the risks of death were significantly decreased but was still significant. After further adjusting for gender (men or women), address (Wuhan city or not), and clinical severity (severe or not), patients with hypertension (HR 1.55, 95%CI 1.35-1.78), diabetes (HR 1.35, 95%CI 1.13-1.62), CBVDs (HR 1.70, 95%CI 1.43-2.02), chronic kidney diseases (HR 2.09, 95%CI 1.47-2.98), and at least two comorbidities (HR 1.84, 95%CI 1.55-2.18) had significant increased risks of death (**Table 3**).

**Table 3.** The associations between comorbidities and the risk of death in patients with COVID-19.

|  |  | HR (95% CI) |  |  |  |
| --- | --- | --- | --- | --- | --- |
|  |  | model1 | model2 | model3 | model4 |
| Hypertension |  | 3.74(3.27,4.28) | 1.7(1.48,1.95) | 1.67(1.46,1.92) | 1.55(1.35,1.78) |
| Diabetes |  | 3.25(2.71,3.89) | 1.66(1.39,1.99) | 1.65(1.37,1.97) | 1.35(1.13,1.62) |
| CBVDs |  | 5.08(4.29,6.02) | 1.98(1.67,2.36) | 1.95(1.64,2.31) | 1.7(1.43,2.02) |
| Lung diseases |  | 2.5(1.89,3.3) | 1.33(1.01,1.75) | 1.26(0.95,1.67) | 1.06(0.8,1.4) |
| Chronic kidney diseases |  | 5.05(3.55,7.18) | 3.25(2.29,4.63) | 3.23(2.27,4.6) | 2.09(1.47,2.98) |
| comorbidities |  |  |  |  |  |
|  | 0 | ref | ref | ref | ref |
|  | 1 | 3.01(2.59,3.5) | 1.56(1.34,1.82) | 1.54(1.32,1.8) | 1.36(1.16,1.58) |
|  | ≥2 | 6.78(5.76,7.98) | 2.48(2.09,2.93) | 2.4(2.03,2.85) | 1.84(1.55,2.18) |

Note: model1: crude model; model2: covariates included age; model3: covariates included age and gender; model4:
covariates included age, gender, address, and clinical severity.

In subgroup analysis (**Table 4**), the association of hypertension, CBVDs, lung diseases, and chronic kidney diseases with the risk of death was similar in both men and women, but the association between diabetes and the risk of death from COVID-19 was prominent in women (HR 1.69, 95%CI 1.27-2.25) than in men (HR 1.16, 95%CI 0.91-1.46). And the interaction effect of gender on the association of diabetes with the risk of death was significant (*P* for interaction = 0.036).

**Table 4.** The associations between comorbidities and the risk of death in patients with COVID-19, stratified by gender.

|  | HR (95% CI) |  | *P* for interaction |
| --- | --- | --- | --- |
|  | Women | Men |  |
| Overall population |  |  |  |
| Hypertension | 1.51(1.2,1.91) | 1.57(1.32,1.86) | 0.944 |
| Diabetes | 1.69(1.27,2.25) | 1.16(0.91,1.46) | 0.036 |
| CBVDs | 1.61(1.21,2.14) | 1.76(1.42,2.19) | 0.791 |
| Lung diseases | 0.94(0.55,1.6) | 1.13(0.81,1.56) | 0.623 |
| Chronic kidney diseases | 1.99(1.12,3.55) | 2.13(1.36,3.33) | 0.997 |
| Comorbidities |  |  | 0.964 |
| 1 | 1.1(0.84,1.43) | 1.51(1.25,1.83) |  |
| ≥2 | 1.81(1.38,2.39) | 1.84(1.48,2.28) |  |

Note: covariates in the model included age, address, and clinical severity.

## 4 Discussion

In the present study, we found that hypertension, diabetes, and CBVDs were the most prevalent comorbidities, and 27.37% of patients with COVID-19 reported having at least one comorbidity. After adjustment for age, gender, address, and clinical severity, COVID-19 patients with hypertension, diabetes, CBVDs, chronic kidney diseases were significantly associated with increased risk of death. And a significant sex-specific difference on the association between diabetes and the risk of death was observed. To the best of our knowledge, this is the largest sample among the reports so far on the association between comorbidities and the risk of death.

Our observations were in line with previous findings in terms of the commonness of comorbidities in patients with COVID-19.^4,5,10-12^ Few studies, however, have explored the association between comorbidities and mortality of SARS-Cov-2 infections. A recent cohort study indicated that hypertension or diabetes were significantly associated with higher risks of the development of acute respiratory distress syndrome (ARDS), but there was no significant correlation between hypertension or diabetes and the risk of progression from ARDS to death.^3^ Guan et al showed that the presence of hypertension and diabetes was respectively associated with 1.58-fold and 1.59-fold increased risk of death in patients with SARS-Cov-2 infection.^4^ Cheng et al showed that kidney disease on admission was associated with an increased risk of in-hospital death.^13^ Zhou et al illustrated that coronary heart disease, diabetes, and hypertension were critical risk factors associated with in-hospital death.^2^ Nonetheless, a major limitation of these researches has been small sample size. Additionally, sex-specific mortality of SARS-Cov-2 infection has been well elucidated, but no studies have pointed out the sex-specific differences on the correlation of comorbidities with mortality. The present study has therefore added further evidence that comorbidities were associated with a greater mortality in patients with COVID-19 based on the large sample size.

The etiology behind the increased risk of death in COVID-19 patients with comorbidities is likely to be multifactorial. Evidence has pointed out that SARS-CoV-2 uses angiotensin converting enzyme 2 (ACE2) as a cell entry receptor.^14,15^ Recent findings have demonstrated that the SARS-CoV-2 cell receptor gene ACE2 was expressed in a wide variety of human tissues, such as small intestine, kidneys, heart, lungs, and adipose tissue.^16^ Therefore, the coronavirus may enter various human cells via ACE2-dependent pathway and cause functional deterioration, leading to the exacerbation of the original comorbidities and subsequently increasing the risk of death. Additionally, cells infected by SARS-CoV-2 produce elevated levels of pro-inflammatory cytokines which may cause immuno-mediated damage to the lungs and other organs, resulting in multi-organ dysfunction.^11,17,18^ Findings from experimental study have illustrated that diabetic mice had a prolonged phase of severe disease and delayed recovery after infection with MERS-CoV.^19^ Therefore, immune dysregulation and prolonged inflammation might be the critical drivers of greater mortality in COVID-19 patients with comorbidities. However, further studies are required to solve this mystery.

Generally, male sex or diabetes comorbidity were related to a greater risk of death of SARS-CoV-2 infection. Anomalously, a significant stronger association between diabetes and the risk of death in COVID-19 patients was observed in females than in males. Estrogen or estrogen receptor might be the key drivers of the sex-specific difference. Population-based studies have shown that lower premenopausal estrogen levels were associated with higher risk of developing diabetes.^20^ The estrogen has been reported to modulate the differentiation, maturation, lifespan, and effector functions of innate immune cells.^21^ Consequently, the decreased levels of estrogen in females with diabetes might be associated with decreases in immune function or increases in inflammatory mediators, resulting in poor clinical outcomes. A cohort study of people with newly diagnosed type 2 diabetes provided indirectly evidence that female sex confers specific vulnerability to the effects of inflammation on adverse health condition.^22^ Additionally, cellular and animal studies have established that G-protein-coupled estrogen receptor is involved in the regulation of inflammation as well as glucose homostasis. Therefore, we speculated that the levels of estrogen or estrogen receptor might play critical roles in the progression of COVID-19 in patients with diabetes. Clearly, experimental studies are needed to elucidated the underlying reasons.

Even though this study included a large number of patients with COVID-19, several limitations should be noted. First, the disease severity and duration of comorbidities, as well as some clinical laboratory indicators (e.g., inflammatory markers) was unavailable in the present study. Second, self-reported diagnosis of comorbidities might result in underestimation of the prevalence of comorbidities, and the existence of recall bias might inevitably affect our assessment. Last, the duration of follow-up was relatively short, and studies with sufficiently long time frame are warranted in future.

## 5 Conclusion

Among laboratory-confirmed cases of COVID-19 in Hubei province, China, patients with hypertension, diabetes, CBVDs, chronic kidney diseases were significantly associated with increased risk of death. The association between diabetes and the risk of death tended to be stronger in women than in men. Clinicians should increase their awareness of the increased risk of death in COVID-19 patients with comorbidities.

## Data Availability

There are no linked research data sets for this submission. The following reason is given:
The authors do not have permission to share data.

## Acknowledgment

We give special thanks to all the healthcare workers in frontline.

---

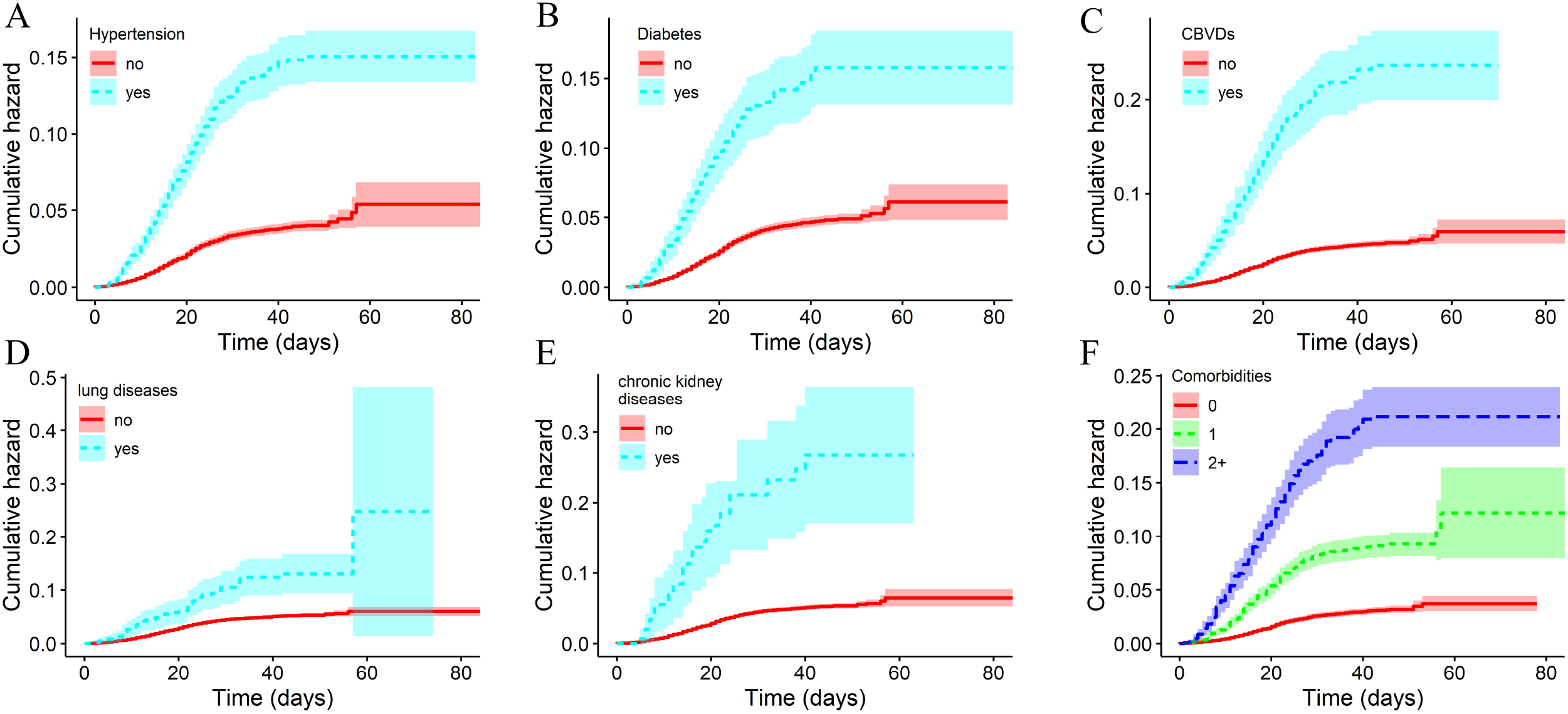

